# The impact of armed conflict on the epidemiological situation of Coronavirus disease (COVID-19) in Libya, Syria, and Yemen

**DOI:** 10.1101/2021.02.12.21251654

**Authors:** Mohamed A Daw

## Abstract

**Background:** Since the Arab uprising 2011, Libya, Syria, and Yemen have gone through a major armed conflict. This resulted in a high rate of mortality, injury, and population displacement with a collapse of the health care system. Furthermore, it was complicated by the emergence of, COVID-19 as a global pandemic which made the population of these countries strive under unusual conditions to tackle both the pandemic and the ongoing wars. The objectives of this study were to determine the impacts and influence of armed conflicts on the epidemiology of Novel Coronavirus (SARS-CoV-2) within these war-torn countries and outline the needed strategies to combat the spread of the pandemic and its upcoming consequences.

**Methods:** The official and public data regarding the dynamics of armed conflict and the spread, of SARS-COV-19 in Libya, Syria, and Yemen were collected from all available sources. Starting from the early emergence of the COVID-19 in each country until the end of December 2020. Datasets were analyzed through a set of statistical techniques and the weekly resolved data were used to probe the link between the intensity levels of the armed conflict and the spread of the pandemic.

**Results:** Data indicated that there is an increase in the intensity of violence levels at an early stage from March to August reached up to two folds in the three countries particularly in Libya. In this violent period, few cases of COVID-19 were reported ranging from 5-53 cases/day. From September to December, a significant decline in the level of the armed conflict was accompanied by steep upsurges in the number of reported COVID-19 cases reached up to 500 cases/day. The highest accumulative cases of COVID-19 were reported in Libya, Syria, and Yemen respectively.

**Conclusions:** Our analysis demonstrates that the armed conflict has provided an opportunity for SARS-COV-19 to spread. At the early weeks of the pandemic that coincided with high levels of the armed conflict few cases were officially reported indicating a vast undercount, which may suggest a hidden mitigating spread at an early stage. Then the pandemic increased immensely as the armed conflict decline to reach the highest by December. A full-blown transmission of the COVID-19 pandemic in these countries is expected. Therefore, urgent national and international strategies should be implemented to combat the pandemic and its upcoming consequences.

## INTRODUCTION

Armed conflicts have major impacts and immense consequences that are difficult to deal with. Historically, one of the most devastating environmental consequences of war is the disruption of peacetime human–microbe relationships, leading to outbreaks of infectious diseases. War produces a multitude of opportunities for pathogenic microbes and constitutes an extremely effective way to promote microbial traffic and increase human morbidity and mortality (1). In the Early Modern Age, when persistent conflicts marred the European continent, the spread of plague (caused by *Yersinia pestis*) was probably aggravated and enhanced through populations fleeing war zones, increasing the geographical range of epidemics(2). During the global pandemic spread of COVID-19, the technically advanced countries focusing on the domestic impact of COVID-19 just as the disease is likely to spread to poor and war-affected countries. Which has the potential to wreak havoc in these fragile states. This is evident in Libya, Syria, and Yemen who have been locked into incredibly destructive armed conflicts for almost a decade now (3). The conflict has resulted in a high rate of mortality, injury, and population displacement. Further to severe destruction of health care system which makes it profoundly ill-prepared for COVID-19 (4).

The oil-rich country of Libya was plunged into chaos after a 2011 NATO-backed uprising toppled and killed the long-standing socialist leader Mummer Gaddafi and split the country into two riving governments(5). In Syria, The first phase of the conflict was ignited by protests in early 2011 what called the Arab Spring uprisings. Then complicated by International sanctions and military intervention of foreign power including Russia, USA, Europe, Turkey, Arab Gulf States & Iran(6).

In Yemen, after the 2011 uprising broke out, the country went through a series of political upheavals and cycles of violence that tore the country apart. Including the start of a full-scale civil war in 2014 and the Saudi- and UAE-led intervention in 2015. This resulted in nearly 100,000 people have died 250,000 displaced and 80% of the population need assistance and protection. Which added further complexity to the COVID-19 pandemic and control measures in such conflict zone (7).

The healthcare system of these three countries at the brink of collapse that alleviates the spread of epidemics. They were the last countries that reported COVID-19 cases in the MENA region. In Syria, the country’s first confirmed case of COVID-19 was reported on March 22, Followed by Libya on March 24, and lately in Yemen on April 10^th^, 2020 (8). These countries are in an extraordinarily poor position to confront the COVID-19 pandemic. Instead of the pandemic leading towards the uniting of local, regional, and international actors involved in these countries around a common purpose, conflict dynamics have hampered an effective response to COVID-19 (9). Furthermore, continued armed conflict would hinder efforts to fight coronavirus and thus act as a catalyzer, and trajectory consequences are expected. However, the influence of armed conflicts on the dynamics of COVID-19 is multifaceted. In this study, we aimed to analyze the impacts and repercussions of the armed conflict on the epidemiological situation of COVID-19 in Libya, Syria, and Yemen and outline the needed strategies to combat COVID-19 and related consequences in these violent regions.

## MATERIALS AND METHODS

### Data Collections

All official and public data regarding the dynamics of armed conflict and the emergence, of SARS-CoV-2 covering the three war-torn countries (Libya, Syria, and Yemen) were collected from the onset of the COVID-19 pandemic till the end of December 2020.

### COVID-19 Dataset

The data for the COVID-19 pandemic were collected from the three countries included in the study. During the period starting from March 2020 till December 2020. This includes the number of total confirmed cases, deaths, and the overall recovered persons from COVID-19. Such data were collected from the official data of the Libyan Ministry of health ((https://ncdc.org.ly/Ar/) and daily reports on its Facebook page about PCR-confirmed infections, deaths, recoveries, and other information. In Syria from Ministry of Health daily COVID-19 updates on their Facebook page (https://www.facebook.com/MinistryOfHealthSYR) and (https://www.worldometers.info/coronavirus/country/syria/). In Yemen(Yemen(https://www.worldometers.info/coronavirus/country/yemen/ Publically available mobility data were collected from; Google (https://www.google.com/covid19/mobility/)which provides data on movement in each country and Government Response Tracker from Oxford Covid-19(https://www.bsg.ox.ac.uk/research/research-projects/coronavirusgovernment) which provide the real-time data on the spread of pandemic worldwide (10). Besides, we used data repository for the 2019 Novel Coronavirus operated by the Johns Hopkins University Center for Systems Science and Engineering (JHU CSSE) to illustrate the unfolding and spatial distribution of conflict events before and during the pandemic and combine this with three brief case studies of Syria, Libya, and Yemen (11).

### Armed conflict Dataset

Armed conflict encompasses all forms of wars and events that cause population death and displacement including civil wars, insurgencies, rebellions, or battles. Each conflict was sorted by date and duration. Furthermore, armed conflict events in Libya, Syria, and Yemen are culled from the Armed Conflict Location &Event Data Project (ACLED). ACLED collects real-time data on the locations, dates, actors, fatalities, and types of all reported political violence and protests events across Africa, Asia, Eastern, and Southeastern Europe, Latin America, and the Caribbean. It provides high quality and widely used source of real-time data on political violence and protests around the world (see https://www.acleddata.com). To avoid misinterpretation, we focus on overall trends, not on individual events that may be poorly expressed in the time series(12).

### Statistical analyses

The collected data were analyzed using Xl-Stat2017 and PAST, version 2.17c. A simple smoother (smoothing transform with moving average as basic function; on weekly bases) was first applied to assess beak trends of the armed conflicts and COVID-19 spread (Accumulative number of cases, and fatalities)(13).

## RESULTS

The armed conflicts and the epidemiological manifestations of the SARS-CoV-2 epidemic in Libya, Syria, and Yemen have gone through various stages and different scenarios. Based on the data available the trajectory of conflicts and the emergence of the global pandemic by March 2020 vary greatly from each of the three countries.

### The Case of Libya

The Libyan armed conflict started in 2011, with the intervention of NATO and toppling the long lasting socialist regime. Since then the country has entered into different upheavals which have been raging in 2014, by splitting control over the country between counterattack governments in east and west of Libya. The East authority had made progress from its self-declared capital in Benghazi, nominally controlling a large portion of the country and, in April 2019, launching a vicious assault on Tripoli, the capital of the Western region government. Figure 1 shows the geographic area controlled by fighting groups. Tripoli and The central area was controlled by the Tripoli authority while the eastern region, Sebha, and most of the Western mountain regions were under the control of East-government. While the southern border region was under the control of other militias. By August 2020, the Tripoli government has rebuffed the eight-month siege of Tripoli and began pushing the Benghazi troops back, reclaiming territory and reasserted control over much of the northwestern corner of the country. This assault has caused over 20,000 deaths, displaced more than 210,000 others, and 1.3 million people in Libya require humanitarian assistance.

**FIGURE 1;.**
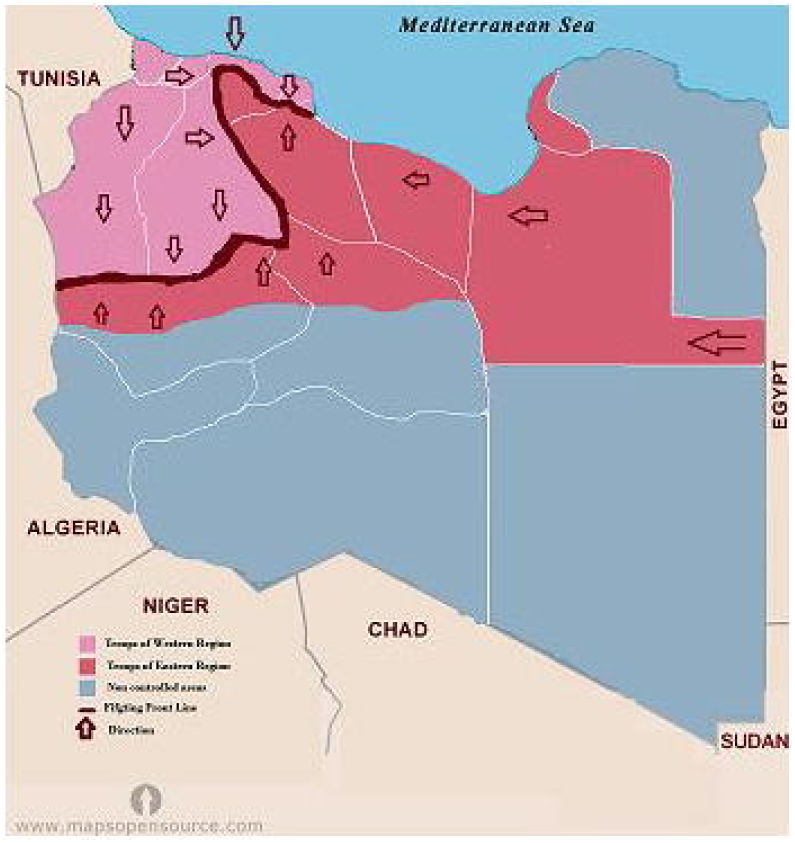
Map of Libya showing the geographic area controlled by each fighting group during the emergence of COVID-19(2020

The arrival of COVID-19 to Libya has coincided with the ignition of the fighting as Figure 2 shows. The conflict continues to rage with an intensification of ground clashes, aerial attacks, and indiscriminate shelling. In March 2020 few cases of COVID-19 were reported in the first eight epi-weeks. Then the reported cases increased significantly to reach over 500 cases/Day by August 2020.

**FIGURE 2;.**
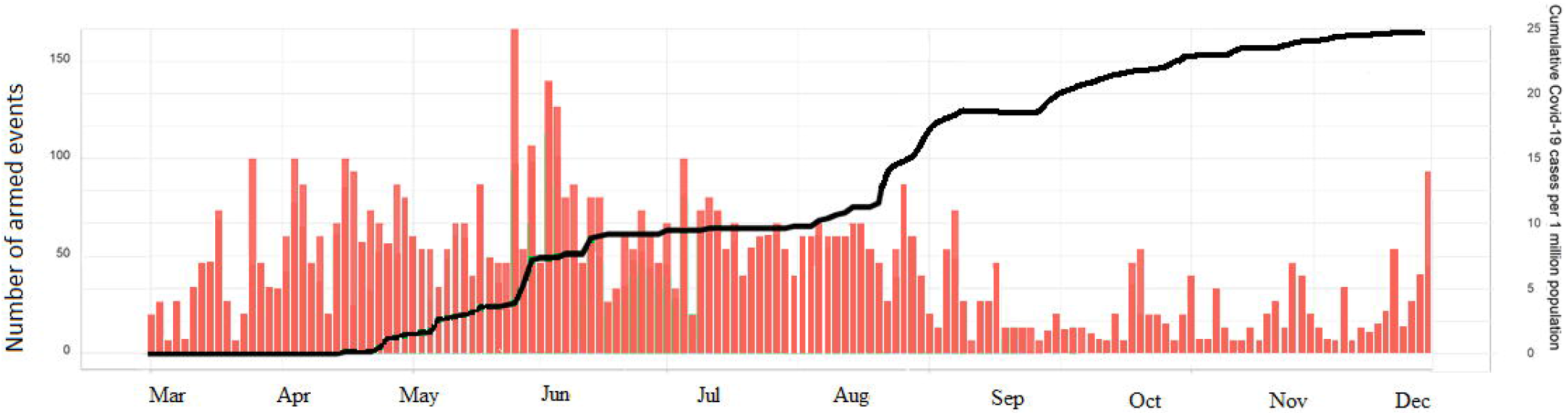
Intensity of the armed conflict and patterns COVID-19 cases in Libya during the epidemic period (March until December 2020)

As of December 2020 (Epi-week 51), a cumulative 95 708 cases has been reported since the first case of the disease was reported on 24 March 2020 as depicted in Figure 3. Of the cumulative number of cases, 28 247 people remain actively infected and 66 076 have recovered. During the reporting period, the cumulative number of deaths rose to 1385. Indicting 1405 cases of COVID-19 per 100 000 population with 20 deaths per 100 000 population with a national case fatality rate (CFR) of 1.4%. The municipalities that have reported large numbers of confirmed **cases** include Tripoli (3874/100 000), Misrata (2061/100 000), and Jabal al Gharbi (1515/100 000). The much lower rates in Sebha (935/100 000) and Benghazi (367/100 000) reflect the low number of tests conducted. The municipalities that have reported large numbers of COVID-19 deaths include Al Kufra (76/100 000), Nalut (52/100 000), and Zwara (43/100 000), which has overtaken Azzawya (42/100 000) from the last situation update. The lower rates in Sebha (30/100 000) and Benghazi (9/100 000) are again a reflection of low levels of case notifications and testing.

**FIGURE 3;.**
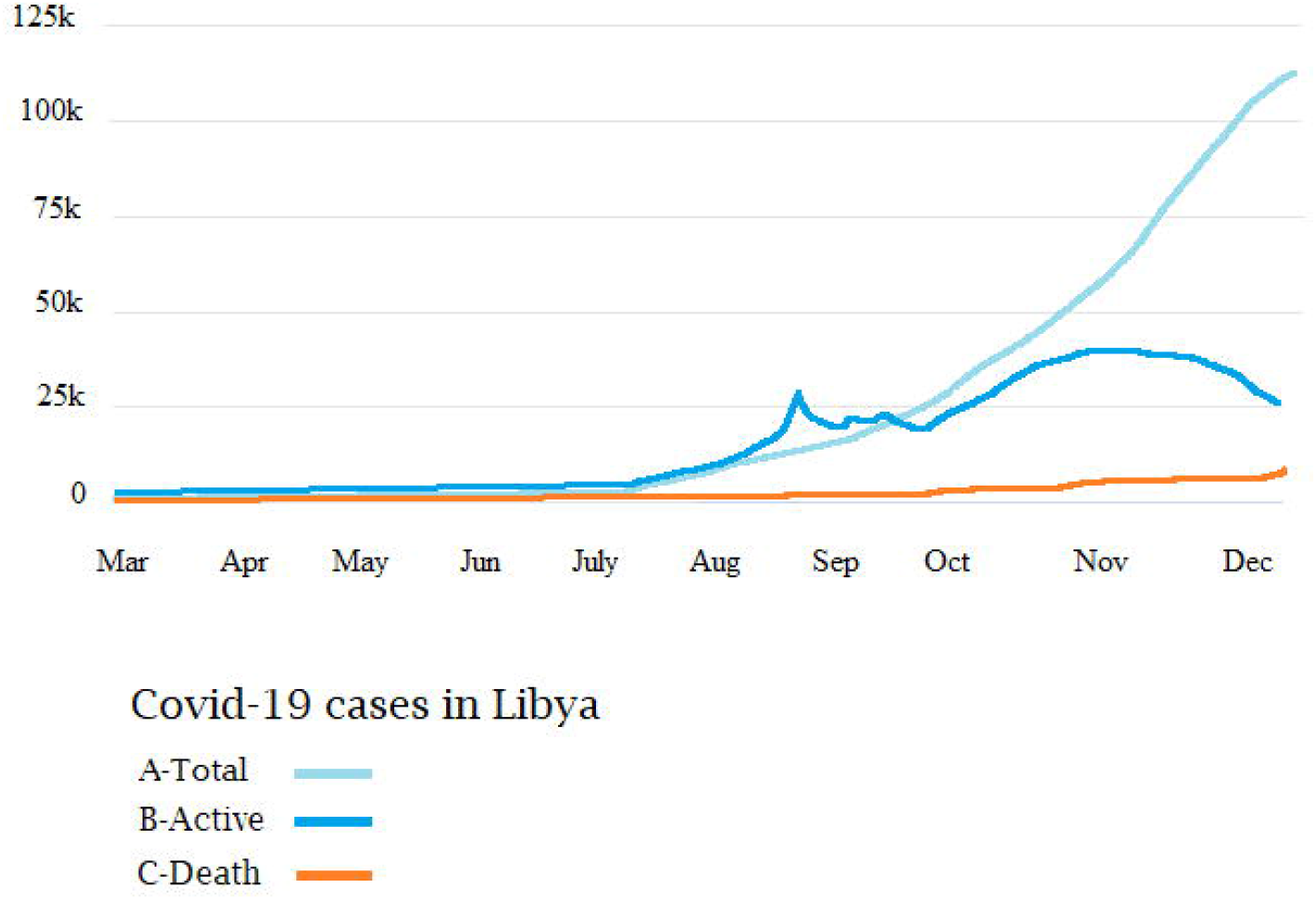
The epidemiological patterns of total cumulative cases (A), active Cases (B), and death cases (C) of COVID-19 in Libya during 2020.

### The Case of Syria

Syria has been plugged, in a decade of armed conflict, which has also decimated Syria’s health system and divided it among several fragmented areas of military control as shown in Figure 4. The country has been torn into five different areas. The armed opposition groups still control large parts of the northern borders including Jarablus, Afrin, Idlib Province, and Hayat Tahrir al-Sham with an estimated 4.2 million people. While the Syrian government has regained control of most of the other parts of the country.

**FIGURE 4;.**
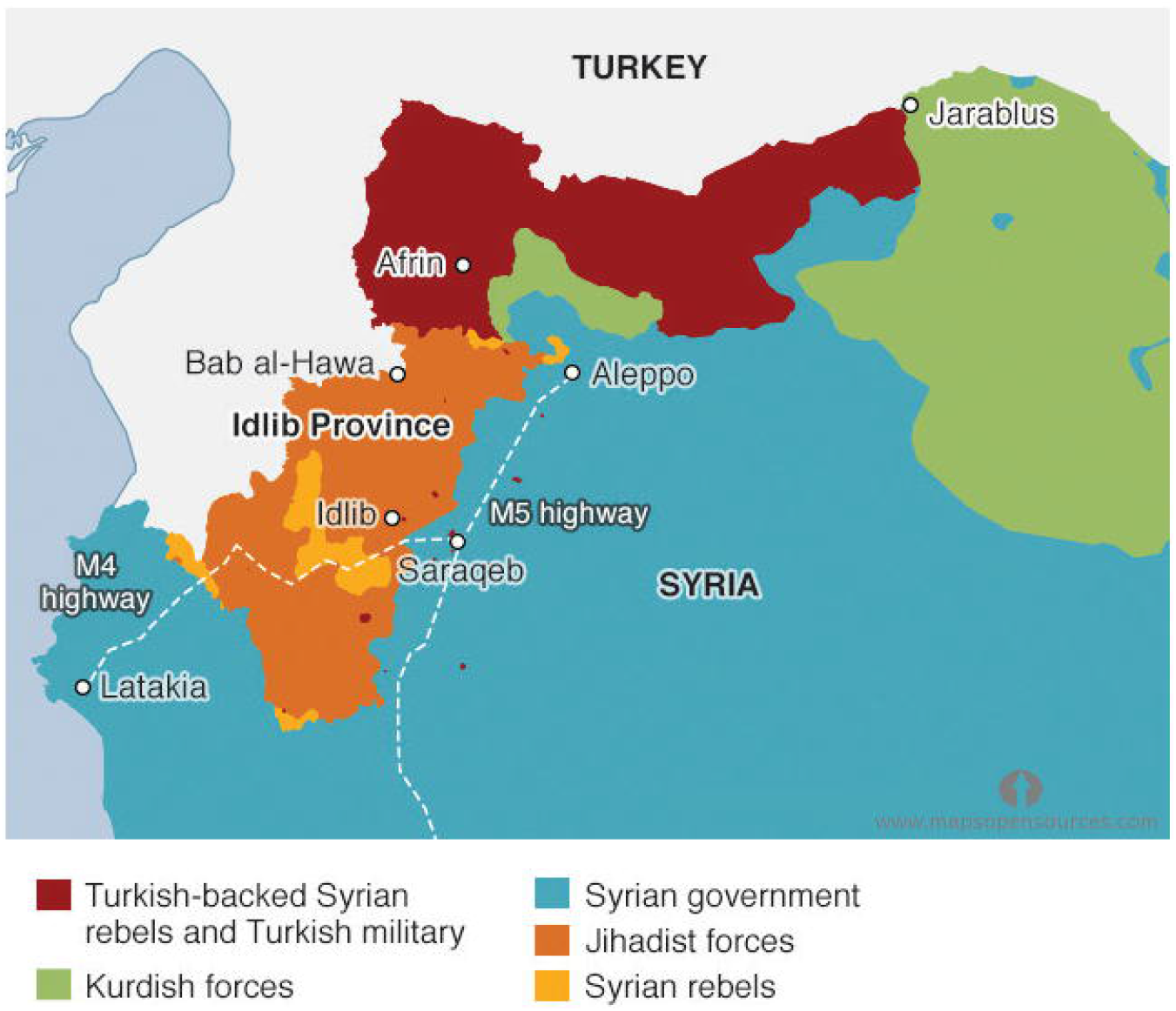
Syria’s map showing geographic areas controlled by each Armed group-2020

In 2020, the Syrian armed conflict has been intensive particularly in Hayat Tahrir al-Sham and Idlib which comes in-concordance with the emergence of the COVID-19 pandemic as shown in Figure 5. The level of violence in Syria remains high and attacks on healthcare Services also appear to have increased. Syria diagnosed its first case of COVID-19 on March 23; for the first two months, the virus spread slowly, never infecting more than ten people per day. By July the number of reported cases started to increase to reach its highest by December 2020.

**FIGURE 5;.**
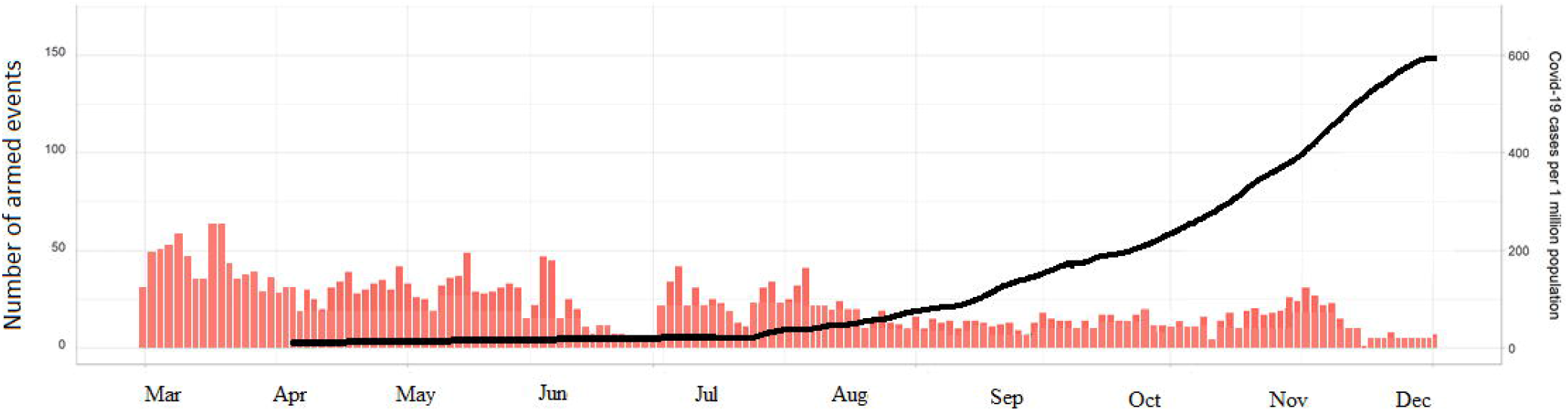
The levels of armed violence in Syria to the spread of COVID-19 during 2020

Figure 6 shows the incidence of accumulative cases of COVID-19 in Syria. A total of 13,885 Coronavirus cases were reported. The prevalence was steady till July 18 th (Ep-week 16) then a slight increase was noticed from August 1^st^ till October 24 ^th^. A sudden increase was noticed form from November 7 till December where the highest cases were reported. During this period 7,329 cases were recovered and only 906 death cases were officially reported. Although the estimated number suggested it may reach 4,340 (95% CI: 3,250 - 5,540) deaths. The highest number was reported in Damascus which was estimated to be 9,760 (95% CI: 6,470 - 11,360) newly infected individuals, including both asymptomatic and symptomatic infections. Further, there is no officially confirmed Covid-19 cases yet from Idlib Jarablus, Afrin, and Hayat Tahrir al-Sham.

**FIGURE 6;.**
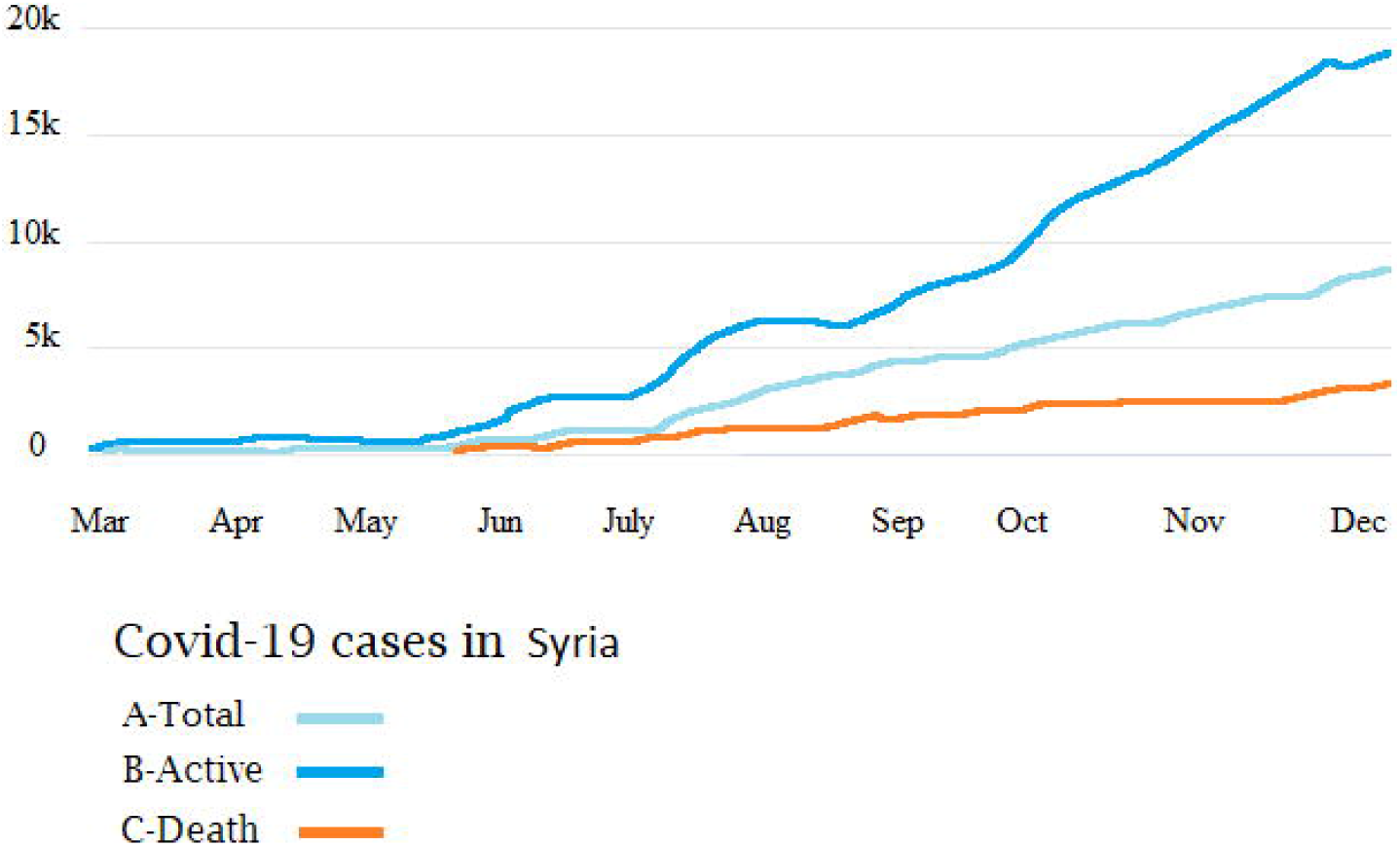
The number of officially reported cases of COVID-19 in Syria, A-Accumulative cases B-Recovered cases C- Number of deaths

### The Case of Yemen

After Yemen’s 2011 uprising broke out, the country went through a series of political upheavals and cycles of violence that tore the country apart, including the start of a full-scale civil war in 2014 and the Saudi- and UAE-led intervention in 2015. The country is now divided into five cantons of political and military control as shown in Figure 7. Including Huthi-controlled northern highlands; Eden government-aligned areas in Marib, al-Jawf, northern Hadramawt, al-Mahra, Shebwa, Abyan, and Taiz city; the pro-separatist Southern Transition Council-dominated territories in Aden and its hinterland; districts along the Red Sea coast where the Joint Resistance Forces are the chief power; and coastal Hadramawt, where local authorities prevail. The war rages along multiple fronts, each with its political dynamics and lines of command and control (Figure7).

**FIGURE 7;.**
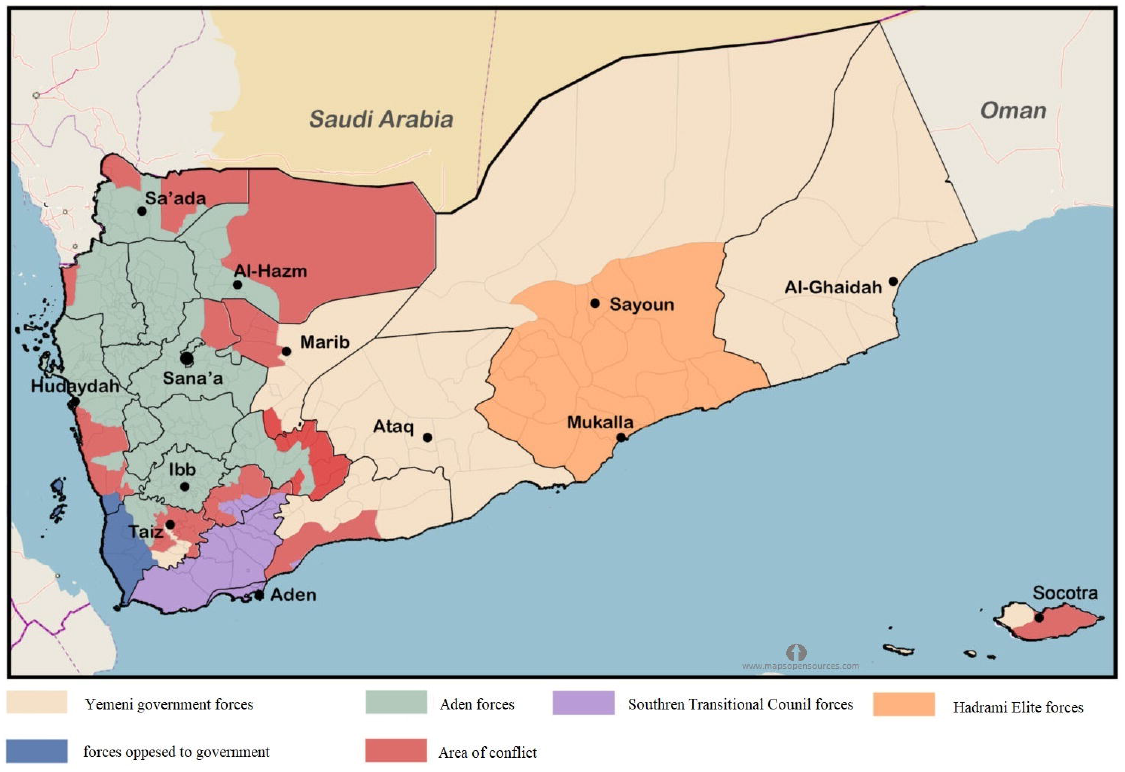
Map of Yemen showing the zones of the armed conflict and areas controlled by each fighting group (2020)

On April 24, 2020, armed confrontations have re-emerged between the Aden army and the forces of the Southern region. These confrontations exhibited further escalation of the armed conflict, which goes parallel with the dissemination of the COVID-19 pandemic within the country in as shown in Figure 8. Yemen announcing the first case on April 10. By the end of May, all of Yemen’s 38 hospitals designated for COVID-19 hospitals were full, yet the Aden government had officially confirmed only 323 cases of coronavirus, including 80 deaths in areas it controls, while Sana authorities had confirmed just four cases, including one death in the whole territory.

**FIGURE 8;.**
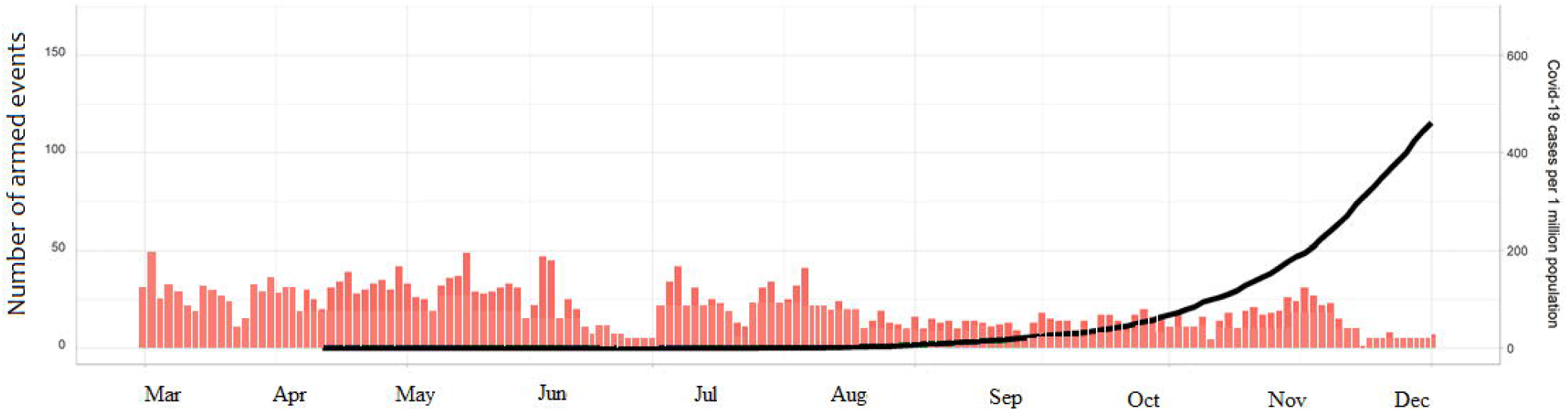
Levels of Violence and COVID-19 cases in Yemen during 2020

Figure 9 shows a total of 2,120 cumulative and 1,425 recovered cases of COVID-19 were reported in Yemen by December 2020. The infection started to rise substantially at the end of April, by August 11, (Epi-week-12) the country has 1,832 confirmed cases and within 26 th September(Epi-week 16) the number of confirmed cases in Yemen had reached 2,034. This was substantially increased by December 31 to reach the highest with a total of 615 deaths. Most of these cases were reported with Eden controlled government.

**FIGURE 9;.**
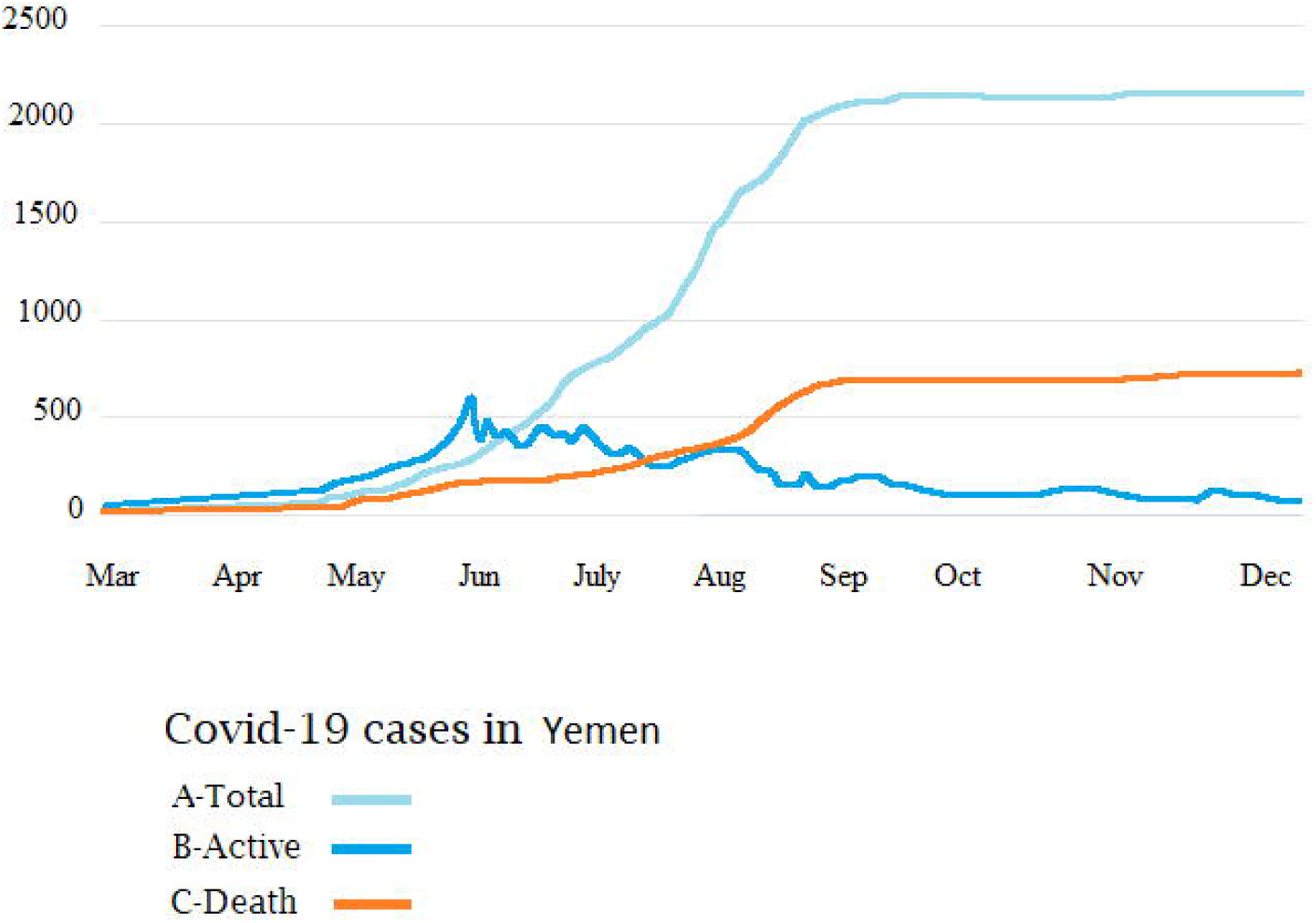
The accumulation levels of COVID-19 cases in Yemen, A-Total cases B-Recovery cases C- Number of deaths

## DISCUSSION

Since its emergence as a pandemic, COVID-19 has affected the whole world and no country can be considered safe. Most of the epidemiological studies were carried in developed and politically stable developing countries. Studies on the impact of armed conflicts on the spread of COVID-19 in conflict hotspots were rarely reported. The effects of armed-conflict on infectious diseases are multi-faceted and complex (14). However, counter-intuitive observations of both increase and decrease in infectious disease outbreaks during and after conflicts have been noticed.

Libya, Syria, and Yemen have experienced one of the most devastating armed conflicts in recent history. Ten years of war have killed, injured, displaced thousands of people, and destroyed the infrastructure and health-care services in these three countries. In Libya, about 1.3 million citizens require humanitarian aid, 200,000 displaced people, and about 636,000 migrants and refugees (5,13). While in Syria About 11.7 million in need of humanitarian aid, and there were about 6.2 million displaced besides 83 percent of Syrians live below the poverty line (15). The conditions are even worse in Yemen as the country witnessing the worst humanitarian crisis in the world, where more than 80% of the population rely on humanitarian aid for survival(16). In light of these poor living-conditions, COVID-19 began spreading in Syria, Libya, and Yemen. It is undoubtedly the case that COVID-19 exacerbated the economic and social problems in these countries due to the lockdown and consequent cessation of many economic activities (17). Hence then, there is an urgent need for quantitative assessment of the effects of the armed conflict on the spread of COVID-19 in war-torn countries.

These three countries were the last countries to report COVID-19 cases in the region. The first case in Syria was reported on March 10, 2020, followed by Libya on March 24 and lately Yemen on April 10(7). At an early stage of the pandemic, few cases were reported in each country and then increased steadily. The conflicts continued at the same pace or become more severe during the pandemic spread particularly in Libya. Even more, most of the conflicting local parties along with the regional and international actors supporting them have viewed COVID-19 as an opportunity to achieve military and political gains (18).

In Libya, only 156 cases were reported in the early months (March-July) of the epidemic which increased drastically up to 1000 cases/day by August-December. This goes parallel to the escalation of the armed conflict, which ignited on April 4, until August 2020. Similar patterns were noticed in Syria and Yemen though the number of confirmed cases in these two countries is less than that in Libya. As of 21 December 2020 (Epi-week 51), Libya occupied 10th place in the list of the ten countries with the highest number of cases in the Eastern Mediterranean region. It occupied 6th place among the top ten countries with the highest number of deaths in the region(19,20). Similar results were reported in Syria and Yemen. A Modelling study carried by Watson *et* al estimated that the burden of COVID-19 in Syria is immense and predicted that a cumulative total of 39.0% (95% CI: 32.5% - 45.0%) of the population had been infected by 2nd September 2020. Assuming that the epidemic has passed its peak in transmission (21). Of the three conflicts Yemen faces the worst COVID-19 outlook, the actual number of cases is likely much higher than the reported numbers. By August, Yemen has 1,832 confirmed cases and 518 deaths.37 This death rate—greater than 28%—is five times higher than the world average (22).

Our data indicate that the armed-conflict has masked the actual status of the epidemic at an early stage. This is in concordance with previous data published by Daw *et al*., which showed that the ongoing-armed armed conflict in these volatile regions has influenced the spread of the pandemic either by masking the actual prevalence in the armed controlling areas and accelerating the spread of the pandemic in the non-controlled region(23).

The data collected from these war-torn countries are almost certainly a vast undercount given the authorities’ chaotic response to the pandemic, especially considering the apparent concealment of cases in Sana-Yemen, Northern Syria, and East of Libya controlled territories. Furthermore, Lack of testing, population displacement, malnutrition, and poverty are the main factors making reliable figures for the incidence of COVID-19 in these countries are hard to come by.

Although this study illustrated the impact of armed conflicts on the spread of COVID-19 in these war-torn countries. A certain limitation has to be emphasized including; the sources of data collection which were based on a certain instance of gray literature that has its methodological limitations and is sometimes subject to interpretation. Further, the data used was not standardized nor validated and it should be interpreted with cautions(24,25). However, intervention programs should be based on the reality that full-blown transmission is ongoing in these countries, which do not have enough resources to respond to the impact coronavirus will have on the population. Therefore, urgent national and international strategies should be implemented to combat the pandemic and its upcoming consequences(26,27).

## CONCLUSION

This study among the first to analyze the impacts of armed conflicts on the epidemiological manifestations of the COVID-19 pandemic. Based on our data analysis, the evidence is emerging that there was a surge of armed conflict and densifying violent attacks in Libya, Syria, and Yemen. Particularly, in the first four to six months during the spread of the COVID-19 pandemic. This resulted in a hidden spread of the pandemic at the early stage, which was exacerbated lately resulting in a high number of infected cases. Suggesting a syndemic interaction between the spread of the coronavirus and the ongoing armed conflict in these countries indicates that the pandemic and its impacts are likely to evolve for years if not dealt with (28,29). Health care systems in these countries face serious challenges; as such, SARS-CoV-2 could spread rapidly through affected populations, particularly among those in the most vulnerable groups. Including injured patients, internally displaced people, prisoners, and immigrants. Hence then global interventions are needed to stop the armed conflict (as have occurred in Libya), protection of health workers and health facilities, and enormous humanitarian support to prepare for an impending crisis (30,31).

## Data Availability

all data are freely available

## ETHICS STATEMENT

No ethical permission is needed.

## AUTHOR CONTRIBUTIONS

MD contributed solely to the conception, design, and statistical analysis. Wrote, corrected, and approved the whole paper,

## DECLARTION OF INTEREST

The author has no conflict of interest in disclosure

## ACKNOWLEDGMENTS

The author would like to thank the immense help and support offered by the Department of Medical Microbiology & Immunology, Faculty of Medicine, University of Tripoli and Libya Study group of COVID-19, and Libyan Study group of HIV/Hepatitis.

